# Machine learning and AI aided tool to differentiate COVID 19 and non-COVID 19 lung CXR

**DOI:** 10.1101/2020.08.18.20175521

**Authors:** Andrio Adwibowo

## Abstract

One of the main challenges in dealing with the current COVID 19 pandemic is how to detect and distinguish between the COVID 19 and non COVID 19 cases. This problem arises since COVID 19 symptoms resemble with other cases. One of the golden standards is by examining the lung using the chest X ray radiograph (CXR). Currently there is growing COVID 19 cases followed by the CXR images waiting to be analyzed and this may outnumber the health capacity. Learning from that current situation and to fulfill the demand for CXRs analysis, a novel solution is required. The tool is expected can detect and distinguish the COVID 19 case lung rely on CXR. Respectively, this study aims to propose the use of AI and machine learning aided tool to distinguish the COVID 19 and non COVID 19 cases based on the CXR lung image. The compared non COVID 19 CXR cases in this study include normal (healthy), influenza A, tuberculosis, and active smoker. The results confirm that the machine learning tool is able to distinguish the COVID 19 CXR lungs based on lung consolidation. Moreover, the tool is also able to recognize an abnormality of COVID 19 lung in the form of patchy ground glass opacity.

To conclude, AI and machine learning may be considered as a detection tool to identify and distinguish between COVID 19 and non COVID 19 cases in particular epidemic areas.

## 1. Introduction

Important approach to deal with the COVID 19 is to find the positive confirmed cases. However, recognizing COVID 19 cases within a community is still becoming a challenge. Related to that, a wide array of COVID 19 medical diagnosis has been developed to support the detection of the COVID 19 cases (Elmousalami and Hassanien 2020). These means of COVID 19 comprehensive medical diagnosis combined with clinical characteristics and radiological diagnosis tools. Clinical characteristics include human temperature monitoring and reverse transcription polymerase chain reaction (RT PCR) (Hutter et al 2019). The temp. monitoring is based on the direct measurement of human body temperature within ranges of 36·5– 38·8°C (Zaheer and Jones 2020). The advantage of body temp. monitoring is it can provide instantaneous data to detect the potential COVID 19. In other hand, RT PCR, which is relying on laboratory technique, is used to test COVID 19 based on blood sample. Nowadays, real time RT PCR test has succeed to confirm the COVID 19 cases worldwide (Feng et al 2020, Huijun et al 2020).

However besides the aforementioned COVID 19 tests above, one of the considerable gold standard test to detect the COVID 19 confirmed cases is using medical imaging. This method is an important and promising candidate to screen for the COVID 19 cases (Lan 2020). A radiological diagnosis includes computed tomography (CT) scans and chest X ray (CXR) radiographs (Molnar 2019). COVID 19 symptoms can be effectively detected using CT or X ray images. Using the chest CT scans, a radiologist can detect the COVID 19 and also the COVID 19 stage in lung, whether it is on recovery or deterioration stages.

CT scans are indeed more sensitive than RT PCR. However, the interpretation of CXR and CT scan is laborious, time-consuming, and requires radiologist expertise and extensive facilities. Unfortunately, an immediate access to such expertise and facilities may not always be available in all clinical settings. These challenges could be addressed by using automated artificial intelligence (AI). This is also important since the appearance of consolidation in lung for COVID 19 cases may have similarity with other diseases like influenza, tuberculosis, or even active smokers. The AI combined with the machine learning can provide accurate early detection to differentiate and diagnose through detection and classification of early lung deterioration signs using CXR imagery. Respectively, this study aims to assess the use of AI and machine learning aided tool to distinguish the COVID 19 and non COVID 19 cases based on the CXR image.

## 2. Methodology

The methodology used in this study is following Hassanien et al. (2020). The first step is visualization of chest X ray (CXR) lung image and enhancing the contrast of the input images by applying a median filter. Second step is applying Otsu objective function to determine multilevel image segmentation. This continues by classifying the COVID 19 infected lung from non infected based on its resemblance probability using the machine learning. The detail of overall procedures and architecture of the COVID 19 classification system is described in figure below.

## 3.Results

The developed machine learning aided tool is able to differentiate and decide which CXR image is having COVID 19 and which is not or having non COVID 19. The ability to detect and differentiate COVID 19 consolidation in lung is represented in detection probability of consolidation resemblances. If the value of resemblance detection is closed to 100% then the system is considered cannot detect and differentiate the presence of COVID 19 related consolidation in lungs.

The Table 1 informs the detection probability of consolidation resemblance values. The table shows that the values are mostly less than 100%. It ranges from 51.09% to 74.30% and confirms that the system is able to detect and distinguish the lung consolidations. The smallest value is observed for the normal lung. While the lung with influenza A lung has the high value.

**Table 1.**
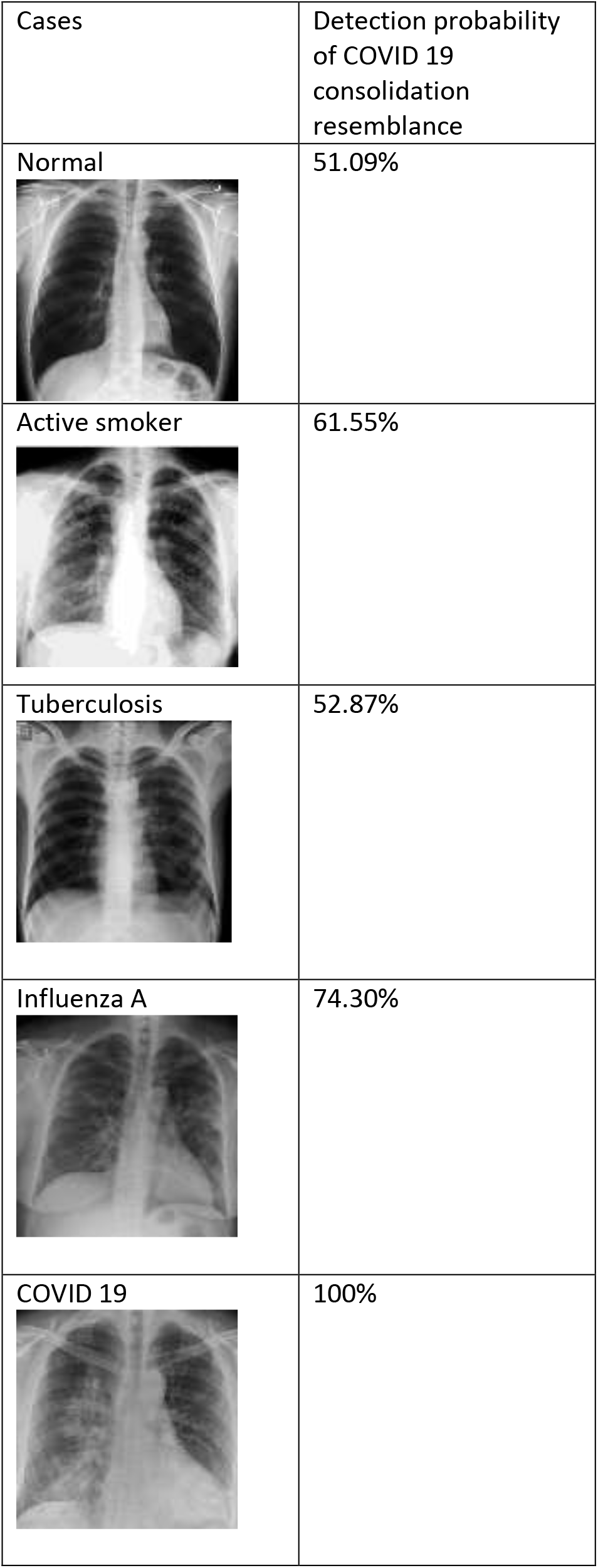
COVID 19 resemblance probability of CXR frontal view image

| Cases | Detection probability of COVID 19 consolidation resemblance |
| --- | --- |
| Normal | 51.09% |
| Active smoker | 61.55% |
| Tuberculosis | 52.87% |
| Influenza A | 74.30% |
| COVID 19 | 100% |

Besides measure the difference between COVID 19 and non COVID 19 lungs, the machine learning is able to detect and mark the abnormalities. Figure 1 shows the abnormality of COVID 19 right lung in the form of patchy ground glass opacity that has been marked. Figure 2 also show the machine learning aided tool can recognize an abnormality as ground glass opacity in left lung.

**Figure 1.**
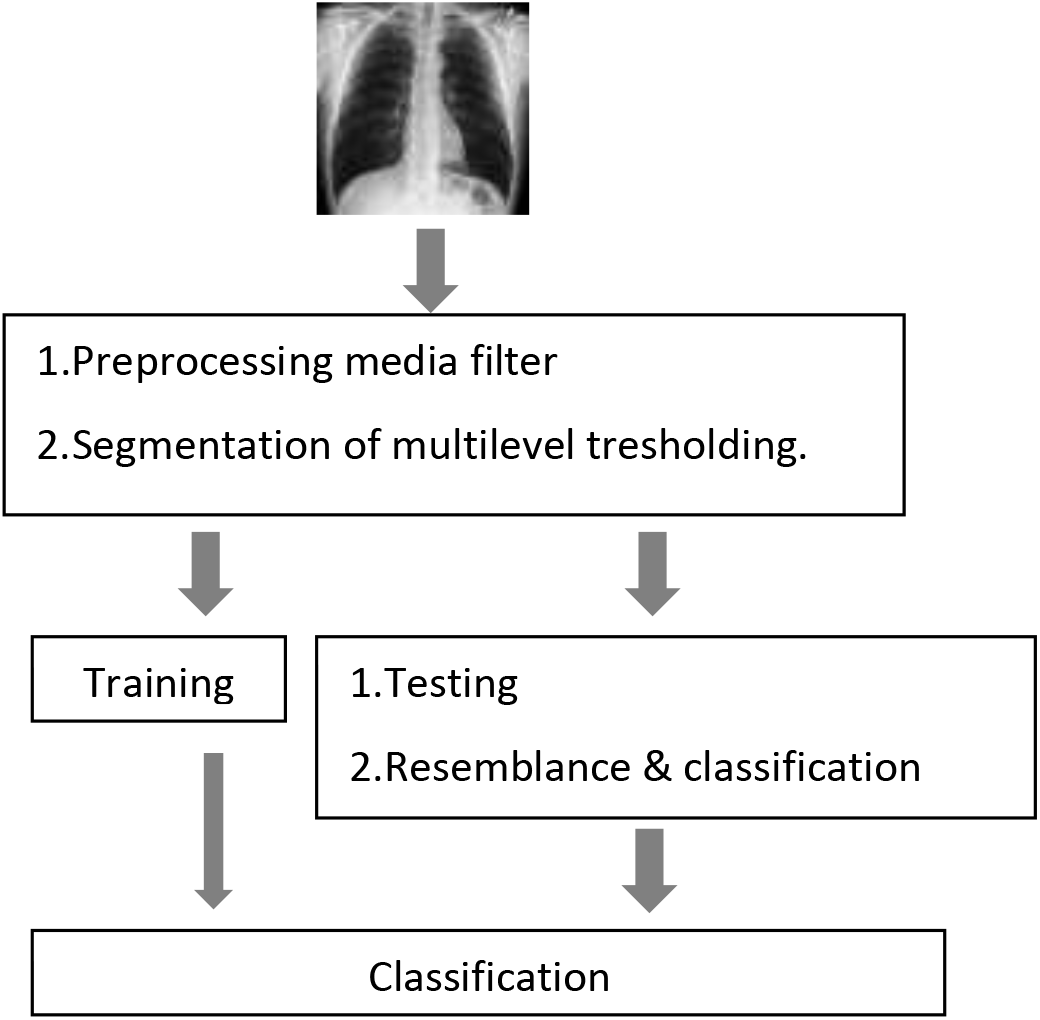
Framework of machine learning aided COVID 19 chest X ray detection(Hassanien et al 2020)

**Figure 1.**
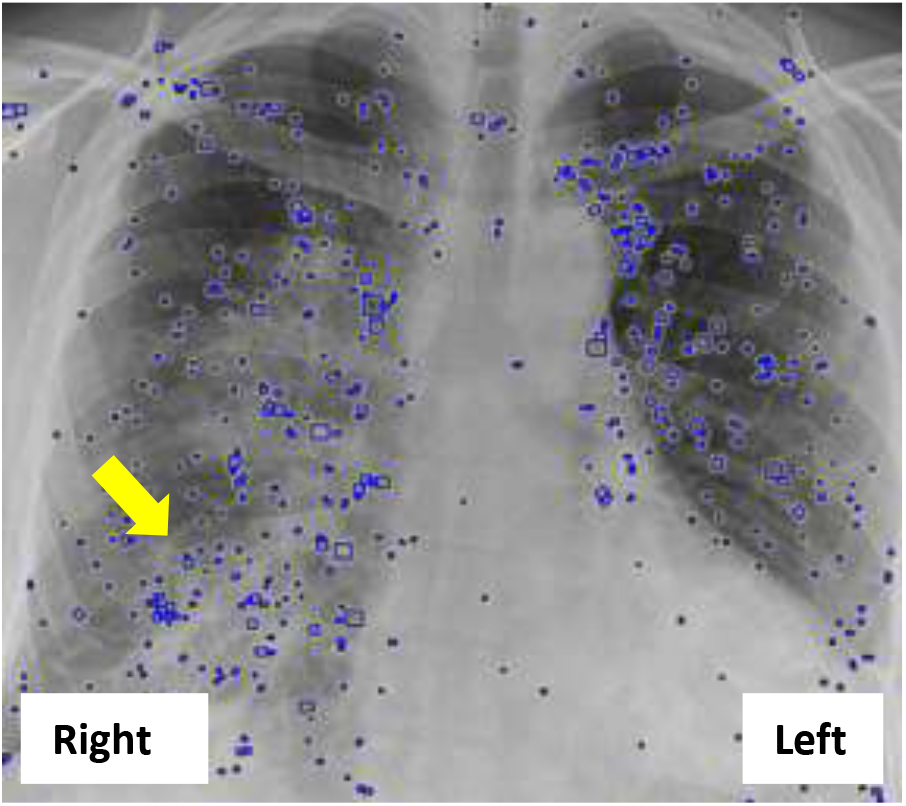
Machine learning detects an abnormality of COVID 19 right lung in the form of patchy ground glass opacity (arrow).

**Figure 2.**
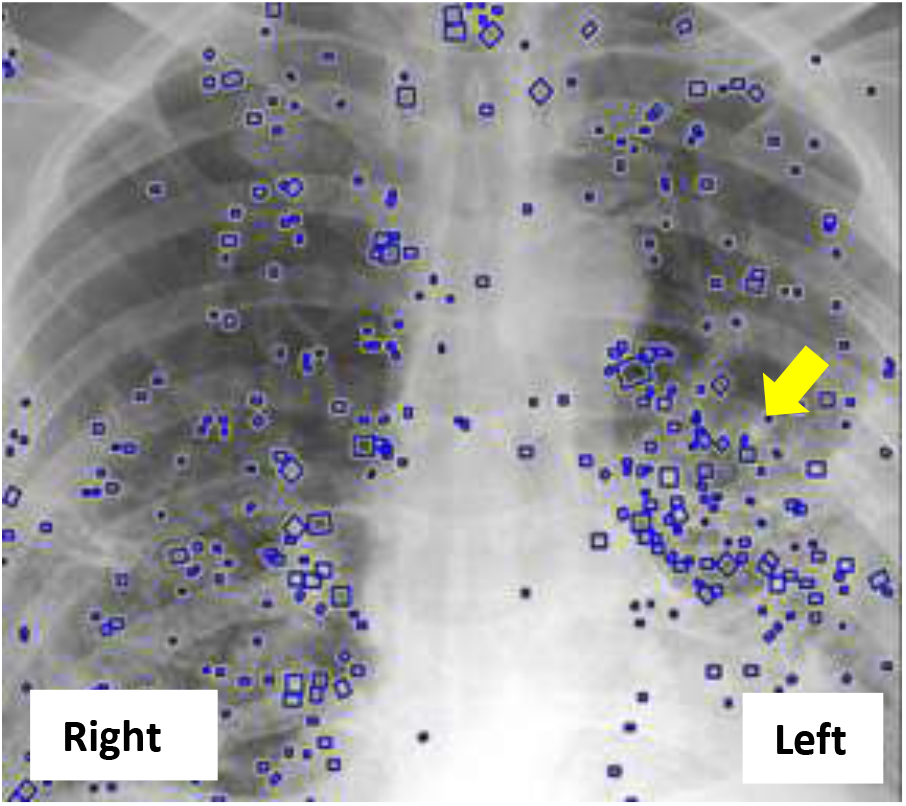
Machine learning detects an abnormality of COVID 19 left lung in the form of patchy ground glass opacity (arrow).

## 4.Discussions

In this study, a CXR of COVID 19 and non COVID 19 cases were used to be analyzed using the AI and machine learning tools. Mangal et al (2020) confirm that the use of CXR has several advantages. First, CXR is more accessible, widespread, and low cost compared to CT scan. In the operational context, CXR is available as portable machine and this enables to conduct X ray test within an isolation ward and reduce the requirement of Personal Protective Equipment. The CXR is also has reliable accuracy to detect COVID 19. Ai et al (2020) reported that sensitivity of CXR was 97% based on positive RT PCR results.

A rapid COVID 19 cases worldwide has demanded a robust and versatile detection tools. Based on the golden standard, the developed tools recently were relied on CXR (Cicero et al 2017) and supported with the analysis performed by AI and machine learning (Chowdhury et al 2020). A lot of studies have been done in this field and noticeable trend of study using AI and machine learning aided medical diagnosis has been observed in the last few years. The trend is growing following recent COVID 19 cases. The result derived from this study is also comparable with the results from those previous studies. A comparison of this study with other reported results is shown in Table 2.

**Table 2.** Current researches of AI used to detect COVID 19 and non COVID 19 CXRs.

| Cases | Author |
| --- | --- |
| Normal, COVID 19 | Castiglioni et al (2020) |
| Normal, COVID 19 | Hassanien et al (2020) |
| Normal, viral pneumonia, COVID 19 | Kana et al (2020) |
| Normal, bacterial pneumonia, viral pneumonia, COVID 19 | Mangal et al (2020) |
| Normal, COVID 19 | Rahimzadeh et al (2020) |
| Normal, influenza, tuberculosis, active smoker, COVID 19 | This study |

The objective of this comparison was to highlight the similarities in performance metrics and effectiveness among previous studies. However, this study has advantage since several non COVID 19 case CXRs have been studied in here as well. As comparison, previous studies (Table 2) have studied only from 2 to 4 non COVID 19 case CXRs. In this study, there are 5 non COVID 19 case CXRs have been studied.

A COVID 19 CXR may resemble with non COVID 19 case CXRs. Then an AI and deep learning tools should have ability and wide coverage to detect multiple cases. It means that the tool besides able to detect the particular COVID 19 CXR, then it should be able to detect and distinguish a lot of non COVID 19 case CXRs without compromise its accuracy.

The growing number of use of AI and machine learning to detect COVID 19 case CXRs related to its advantage in comparison to conventional tools. Hassanien et al (2020) noted that the tool is proven useful to detect COVID 19 CXR. The tool can deliver high performance amid accuracy and computational complexity.

Numerous studies have reported the performance of machine learning tools to detect the COVID 19 in term of sensitivity, specificity, and accuracy. Study by Kana et al (2020) reported the observed accuracy rate, recall, precision, and F score for COVID 19 and non COVID 19 CXRs (bacterial and viral pneumonia, normal) were 95-100%, 98-100%, 97-100%, and 97-100% respectively. While other studies reported performance value ranges equal to 82.8%-98.7%, 91.53%-100%, 94.98%-97.3%, and 92.78%-98.63% for accuracy, precision, recall, and F measure (Cicero et al 2017, Jin et al 2017, Lakhani and Sundaram 2019). Therefore, these evidence based findings demonstrate the reliability of deep learning and AI uses to aid COVID 19 detection.

## 5. Conclusions

The COVID 19 is confusing considering it resembles to other disease include common flu and tuberculosis as examples. This condition leads to confusion about who can, or should, get tested. To deal with this challenge, this study has used an AI and machine learning aided assessment tool to determine whether a person is infected with COVID 19 or not based on the CXR lung assessment.

## 6. Recommendations

Amid ongoing COVID 19 pandemic, a numerous routines include isolation and social distance seem to be temporary unpractical solution. While, COVID 19 vaccine is expected to take at least 18 months only if it works at all. Unfortunately, COVID 19 can mutate and become more aggressive. Learning from this aforementioned situations, then it is recommended to consider the use of AI and machine learning aided COVID 19 detection as one of the solution for the COVID 19 CXR analysis.

Following COVID 19 cases, the demand for CXR will grow exponentially and resulted in the large number of CRX images waiting to be analyzed. Considering that the CRX image analysis may outnumber the capacity of all clinical settings, then applying AI and machine learning aided decision supports are considered to be very useful to support the frontline health practitioners.

## Data Availability

covid chestxray dataset master (ieee8023/covid-chestxray-dataset)

